# A weighted quantile sum regression with penalized weights and two indices

**DOI:** 10.1101/2022.12.10.22283301

**Authors:** Stefano Renzetti, Chris Gennings, Stefano Calza

**Affiliations:** Department of Medical and Surgical Specialties, Radiological Sciences and Public Health, Università degli Studi di Brescia, viale Europa 11, 25123 Brescia, Italy; Department of Environmental Medicine and Public Health, Icahn School of Medicine at Mount Sinai, 17 E 102nd Street, New York, NY 10029, USA; Department of Molecular and Translational Medicine, Università degli Studi di Brescia, viale Europa 11, 25123 Brescia, Italy

**Author notes:** Corresponding Author: Stefano Renzetti, Department of Medical and Surgical Specialties, Radiological Sciences and Public Health, Università degli Studi di Brescia, viale Europa 11, 25123 Brescia, Italy.

**Keywords:** Environmental mixture, weighted quantile sum regression, two indices, penalized weights, nutrients, obesity

## Abstract

**Background:** New statistical methodologies were developed in the last decade to face the challenges of estimating the effects of exposure to multiple chemicals. Weighted Quantile Sum (WQS) regression is a recent statistical method that allows estimating a mixture effect associated with a specific health effect and identifying the components that characterize the mixture effect.

**Objectives:** In this study, we propose an extension of WQS regression that estimates two mixture effects of chemicals on a health outcome in the same model through the inclusion of two indices with the introduction of a penalization term.

**Methods:** To evaluate the performance of this new model we performed both a simulation study and a real case study where we assessed the effects of nutrients on obesity among adults using the National Health and Nutrition Examination Survey (NHANES) data.

**Results:** The method showed good performance in estimating both the regression parameter and the weights associated with the single elements when the penalized term was set equal to the magnitude of the Akaike information criterion of the unpenalized WQS regression. The two indices further helped to give a better estimate of the parameters (Positive direction Median Error (PME): 0.017; Negative direction Median Error (NME): -0.023) compared to the standard WQS (PME: -0.141; NME: 0.078). In the case study, WQS with two indices was able to find a significant effect of nutrients on obesity in both directions identifying caffeine and magnesium as the main actors in the positive and negative association respectively.

**Discussion:** Through this work, we introduced an extension of the WQS regression that showed the possibility to improve the accuracy of the parameter estimates when considering a mixture of elements that can have both a protective and a harmful effect on the outcome; and the advantage of adding a penalization term when estimating the weights.

## Introduction

Humans are exposed to many chemicals from multiple chemical classes, based on biomonitoring data, which may influence a particular disease or health state. (Carlin et al., 2013; Carpenter et al., 2002; Patel, 2017). Of particular concern is that many of these exposures may act jointly (e.g., along a common adverse pathway) so that even low levels of each may together have an adverse effect. Not accounting for this *mixture effect* may underestimate the potential health risk. Analogously, the food we eat includes many important nutrients. Evaluating dietary quality based on a single nutrient is not adequate.

Several new statistical methodologies were developed to face the challenges of estimating the effects of exposure to multiple chemicals, each addressing its own research question (Stafoggia et al., 2017). These new methods were developed to solve problems like multiple comparisons and multicollinearity that are commonly encountered with high dimensional and correlated exposures. When using classical statistical models, an increased probability of incurring false positive or false negative results can occur in the first case (Braun et al., 2016; Bretz et al., 2005; Greenland, 2008; Savitz and Olshan, 1995; Stacey and Czyz, 2012) while multicollinearity can produce unstable and biased parameter and standard error estimates in the second case (Billionnet et al., 2012; Dominici et al., 2010; Dormann et al., 2013; Jain et al., 2018; Leal et al., 2012; Patel, 2017; Stafoggia et al., 2017; Sun et al., 2013; Tu et al., 2008; Vatcheva et al., 2016; Weisskopf et al., 2018).

Weighted Quantile Sum (WQS) regression is a recent statistical method that is increasingly applied in epidemiological studies to address the research questions of (i) is there a mixture effect associated with a specific developmental or health effect; and (ii) which of the measured components characterize the mixture effect. This method builds an empirical weighted index that represents the mixture effect in an ensemble first step and tests its association with the outcome of interest in a second step. This body burden index reduces the dimensionality and is more robust to multicollinearity (Carrico et al., 2015; Czarnota et al., 2015). The original methodology provides the estimate of a single empirically-weighted index that measures the association between the mixture and the dependent variable in only one direction (either positive or negative), which may be interpreted as the joint action of the components in that direction (i.e., a mixture effect). On the other hand, other methods (e.g., shrinkage methods) focus on parsimony in predicting an outcome and thus address different research questions. Further, shrinkage methods suffer from limitations in the presence of highly correlated variables like the grouping effect in the elastic net and the arbitrary selection of variables in the LASSO that can be problematic in the risk evaluation of environmental mixtures (Carrico et al., 2015). The estimate of a single index can be an advantage when the elements in the mixture have the same direction in the association with the dependent variable, as is the case when evaluating a mixture effect of jointly acting components. In fact, looking in one direction we avoid the reversal paradox (Tu et al., 2008) and we increase the power to detect a mixture effect using a single degree of freedom test. However, when the mixture is made of both “good” and “bad” actors (i.e., two sets of components where one set is related to a positive direction and opposite for the other set), it can become a limitation when we want to estimate both the positive mixture effect and negative mixture effect on the specific outcome of interest in a single analysis.

In this study, we propose an extension of WQS regression where two indices are estimated (termed two-indices WQS (2iWQS) in the sequel), one in the positive and the other in the negative direction, in the same model both at the nonlinear estimation ensemble step where the weights are determined and at the final model inference step. This will allow us to use a data reduction approach trying to focus the inference for joint action in now both directions still based on the idea of a mixture effect with a single degree of freedom test for each direction. The simultaneous estimation of the two indices is made possible through the addition of a penalization term when estimating the weights. An application of the new method uses National Health and Nutrition Examination Survey (NHANES) (2015-2016) dietary data and the effect on obesity. A simulation study is also presented to evaluate the performance of the method.

## Methods

The general formula for the WQS generalized nonlinear regression model for *c* components in the mixture is the following:

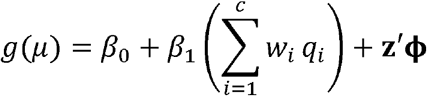

where *w*_*i*_ are the unknown weights associated to each component of the mixture to be estimated through an ensemble step using bootstrap samples or random subset samples, *q*_*i*_ are the values of the components scored into quantiles (quartiles, deciles,…), *β*_0_ is the intercept, *β*_1_ is the coefficient associated to the WQS index, **z′ ϕ** are the vector of covariates and parameters, respectively and *g*() is any link function between the mean *μ* and the predictor variables as in generalized linear models. WQS regression requires that data are split in a training and validation dataset. The first part of the data is used for the ensemble step to determine the weighted index, while the final model is fitted on the holdout data to test the significance of *β*_1_. For the estimated weights the following constraints are imposed: 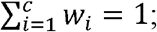 and 0 ≤ *w*_*i*_ ≤ 1.

Once the weights are estimated for each ensemble step sample the WQS index is estimated through the formula: 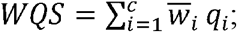 where 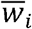 is the mean of the weights found in the ensemble steps associated to either a positive or a negative 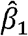 depending on the chosen direction of the association between the mixture and the outcome:

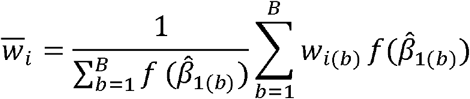

where, for example, 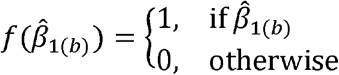 and the chosen direction have the same sign. Other signal functions include the inverse of the absolute value of the t-test statistic for 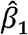, or *t*^2^, or *e*^*t*^. The final model is fitted on the holdout validation dataset using the generalized linear model *g*(*μ*) = *β*_0_ + *β*_1_ *WQS* + **z′ ϕ**.

In this study we propose to first include a penalized weight estimate to better identify the truly associated elements in the case of highly correlated data. The objective function (with an identify link) in this case is as follow:

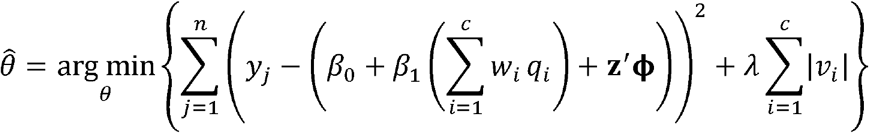

where *θ* = (*β*_0_, *β*_1_, *ν*_1_, …, *ν*_*c*_, **ϕ**, λ) and 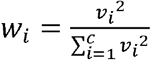, *ν*_*i*_ ∈ ℜ, *i* = 1, … *c*.

We then introduce two indices in the same model to allow an estimate of the mixture effect in a positive direction and a mixture effect in a negative direction at the same time, both in the training and validation steps. The new general formula is the following:

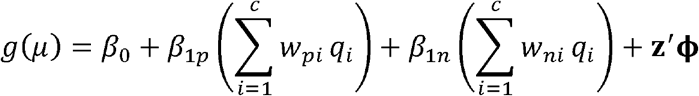

where *w*_*pi*_ and *w*_*ni*_ are the unknown weights associated with each component for the positive and negative direction, respectively, while *β*_1*p*_ and *β*_1*n*_ are the two parameters that measure the positive and negative effect of the mixture on the outcome. The two indices will be kept in the model both in the first step where two set of weights are estimated (one for the positive and one for the negative direction) and in the second step when the final model is fitted. The equality and inequality constraints are applied to both sets of weights besides a constraint to each *β*_1*j*_ parameter: *β*_1*p*_ ≥ 0 and *β*_1*n*_ ≥ 0. The penalization term is also considered to better discriminate between the elements having an effect and those not associated with the outcome and to reduce the noise produced by the null components that can increase the correlation between the two indices. The objective function is of the form:

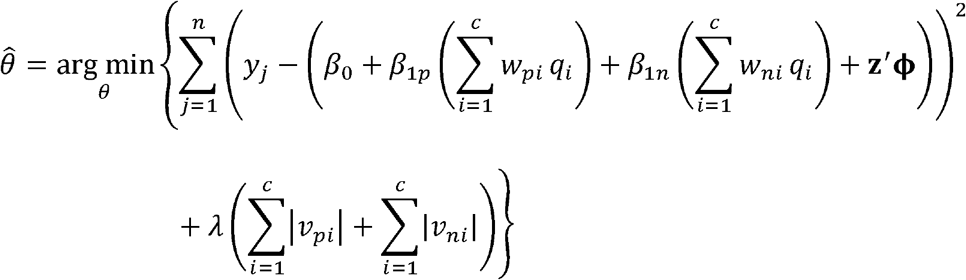

where θ = (*β*_0_, *β*_1*p*_, *β*_1*n*_, *ν*_*p*1_, …, *ν*_*pc*_, *ν*_*n*1_, …, *ν*_*nc*_, **ϕ**, *λ*) and 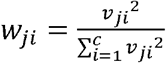, *ν*_*ij*_ ∈ ℜ, *j* = *p, n, i* = 1, … *c*.

To set the starting values of the *ν*_*ji*_ we fit two standard WQS regressions constraining the beta in each direction and running only one bootstrap iteration. We then extract the estimated *ν*_*ji*_ from the two models.

The final model for the validation step is *g*(*μ*) = *β*_0_ + *β*_1*p*_ *WQS*_*p*_ + *β*_1*n*_ *WQS*_*n*_ + **z′ ϕ**.

To further control the collinearity between the two indices we apply a different signal function when averaging the weights estimated in each ensemble sample. In particular, we considered the tolerance (the inverse of the variance inflation factor (VIF)) as the weight in the weighted mean: 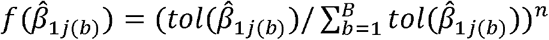 where *tol* is the tolerance associated to the parameter 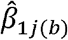, *j* refers to the direction (*j* = *p* for positive direction, *j* = *n* for negative direction) and *n* is chosen depending on the variability of the tolerance values: higher *n* is applied with lower variability to better discriminate between those models where there is higher collinearity between the positive and the negative index and those where collinearity is less severe. For example, we start with *n* = 2; if the VIF for either WQS parameter exceeds 5 then we may increase *n* to, say, 3.

Through this work, we also wanted to provide an outline of the steps to take when trying to fit a WQS regression with a single or double index. To define the shrinkage parameter a cross-validation step should be performed. However, since this can be computationally intense, a rule of thumb that can be applied to choose the value of the parameter lambda is to set it equal to the magnitude of the Akaike Information Criterion (AIC) of the non-penalized WQS regression. We then suggest to fit a non-penalized WQS regression and then run the same model setting three different shrinkage parameter values: one equal to the magnitude of the AIC of the previous regression, one to a lower and one to a greater order of magnitude. In the case of the 2iWQS we suggest to do a final check of the collinearity between the two indices. To apply a WQS regression we then suggest to follow these three steps:

- Step 1: fit a standard WQS regression (with single or double index as needed)
- Step 2: set three different shrinkage parameter values, one equal to the magnitude of the AIC of the regression fitted at step 1 (see the results section, figure 2), one to a lower and one to a greater order of magnitude and follow a rough bisection algorithm. Alternatively, the search of the best lambda can also be refined by looking to the intermediate values, e.g. if the magnitude of the AIC is 1,000, then lambda can be set equal to 500 and 5,000 besides 100 and 10,000.
- Step 3: In the case of the 2iWQS a final check of the correlation between the two indices is needed. The exponent in the signal function can be increased in the case of multicollinearity.

The possibility to fit a 2iWQS and to estimate penalized weights will be added to the gWQS R package (Renzetti et al.).

### Simulation Study

To evaluate the performance of this new model in terms of the estimation of the regression parameters and the weights, we performed a simulation study. We took the data from the NHANES 2011-2012, 2013-2014 and 2015-2016 survey cycles where a total of 38 nutrients were measured through the administration of a food frequency questionnaire. In total 100 different datasets were built generating the 38 variables from a multivariate normal distribution keeping the same correlation structure of the original data. As we can see from figure 1 the nutrient data show a complex correlation matrix ranging from -0.05 to 0.93.

**Figure 1.**
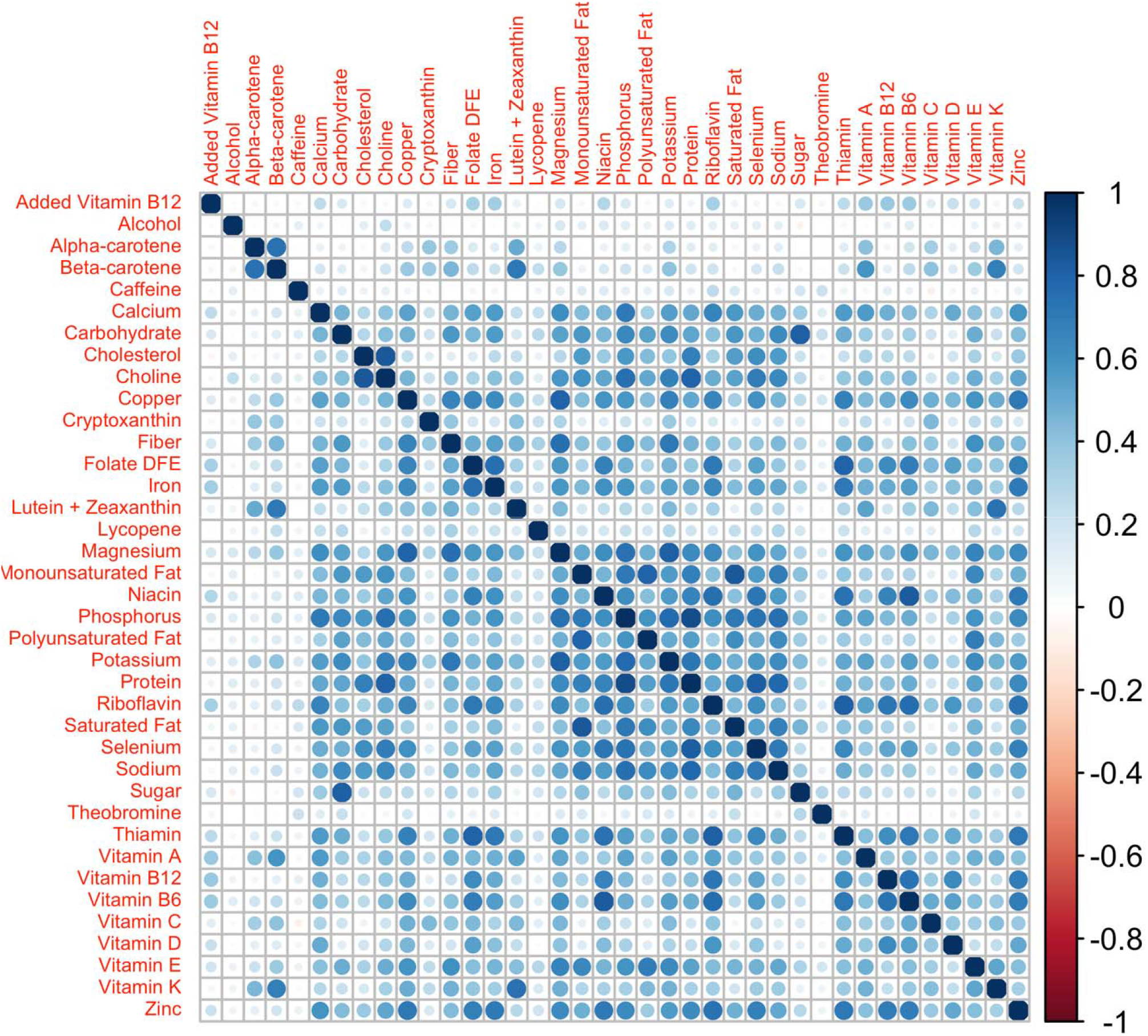
Nutrients’ correlation matrix: Correlation matrix among the 38 nutrients from the NHANES 2011-2012, 2013-2014 and 2015-2016 survey cycles.

Based on the case study results (see the results section, figure 6) the corresponding four nutrients with a median weight exceeding the threshold were selected for the positive direction while the corresponding 8 elements were chosen in the negative direction. In particular, the dependent variable was generated from a normal distribution with mean equal to the combination obtained by applying the parameters given in Table 1 to the 2iWQS formula and a unit standard deviation. The case study weights were rescaled assigning a null weight to all the non-selected nutrients. One more scenario was considered halving the values of the correlation matrix. This simulation study is structured such that the sensitivity in the positive direction (i.e., estimating weights exceeding the threshold of 1/38 = 0.026) is particularly difficult when two of the four nutrients have “true” weights of 0.04 and 0.07.

**Table 1.** Parameters’ value used in the simulation study: Values of the parameter regression and weights used to generate the dependent variable. The weight of all the remaining variables not included in the table were set to 0.

| Parameter | PWQS | NWQS |
| --- | --- | --- |
| $\beta_{1p}$ | 0.5 | |
| $\beta_{1n}$ | | -0.5 |
| $w_{caffeine}$ | 0.71 | 0 |
| $w_{sodium}$ | 0.18 | 0 |
| $w_{polyunsaturated\ fat}$ | 0.07 | 0 |
| $w_{calcium}$ | 0.04 | 0 |
| $w_{magnesium}$ | 0 | 0.4 |
| $w_{beta-carotene}$ | 0 | 0.12 |
| $w_{folate\ DFE}$ | 0 | 0.11 |
| $w_{vitamin\ B6}$ | 0 | 0.1 |
| $w_{fiber}$ | 0 | 0.07 |
| $w_{vitamin\ C}$ | 0 | 0.07 |
| $w_{vitamin\ D}$ | 0 | 0.07 |
| $w_{vitamin\ K}$ | 0 | 0.06 |

Abbreviations: DFE, Dietary Folate Equivalents; PWQS, Positive Weighted Quantile Sum index;
NWQS, Negative Weighted Quantile Sum index.

### Case Study

We then applied the new method of the 2iWQS regression in a real case study. We considered the nutrition information from the NHANES 2011-2012, 2013-2014 and 2015-2016 study cycles and we assessed the effects of nutrients on obesity among adults.

Obesity was defined as Body Mass Index (BMI) greater or equal to 30 kg/m^2^. We excluded subjects with severe obesity (BMI ≥ 40 kg/m^2^).

Nutrients were estimated from the dietary intake data that considered the types and amounts of foods and beverages (including all types of water) consumed during the 24-hour period prior to the interview (midnight to midnight). Two interviews were performed: the first one was collected in-person while the second interview was collected by telephone 3 to 10 days later. Details of the survey are described elsewhere (2016a; 2016b). In this study, we averaged the two nutrients when both evaluations were considered as usual food consumption (only one measurement was included in the analysis if the other one was not usual while the observation was dropped if both evaluations were not usual; this was a self-reported answer to the question of whether the person’s overall intake on the previous day was much more than usual, usual or much less than usual) and we added the dietary supplement intake when applicable.

The models were adjusted by covariates including age, sex, race, education as the highest grade or level of school completed or the highest degree received, the ratio of family income to poverty (using the Department of Health and Human Services poverty guidelines), the minutes of sedentary activity represented by the time spent sitting on a typical day and the minutes of moderate and vigorous activities spent either during work or during recreational activities categorized using its tertiles (because of the skewed distribution), the smoking status as never-smokers (subjects who did not smoke as many as 100 cigarettes in their lifetime), former smokers (those who smoked at least 100 cigarettes in their lifetime but were not currently smoking cigarettes), and current smokers (subjects that currently smoked cigarettes) and the study cycle. Exclusion criteria included being on any kind of diet to lose weight or for another health-related reason at the time of the interview, having a BMI greater than 40 kg/m^2^ and being younger than 20 or older than 60 years old. In total, 5105 subjects were included in the study.

The Kruskal-Wallis test and Chi-squared test were used to test differences between obese and non-obese participants for continuous and categorical variables respectively.

Consent from subjects participating in the study was received prior to conducting the study and the study has been reviewed and approved by the CDC/NCHS Ethics Review Board (ERB).

## Results

### Simulation Study

As a first step we tested for the best shrinkage parameter *λ*: a WQS regression was fitted on each dataset letting *λ* vary among the values 0, 0.1, 1, …, 10^5^. The best shrinkage parameter was selected which minimized the AIC. In our case, *λ* = 100 was the optimum value as shown in figure 2.

**Figure 2.**
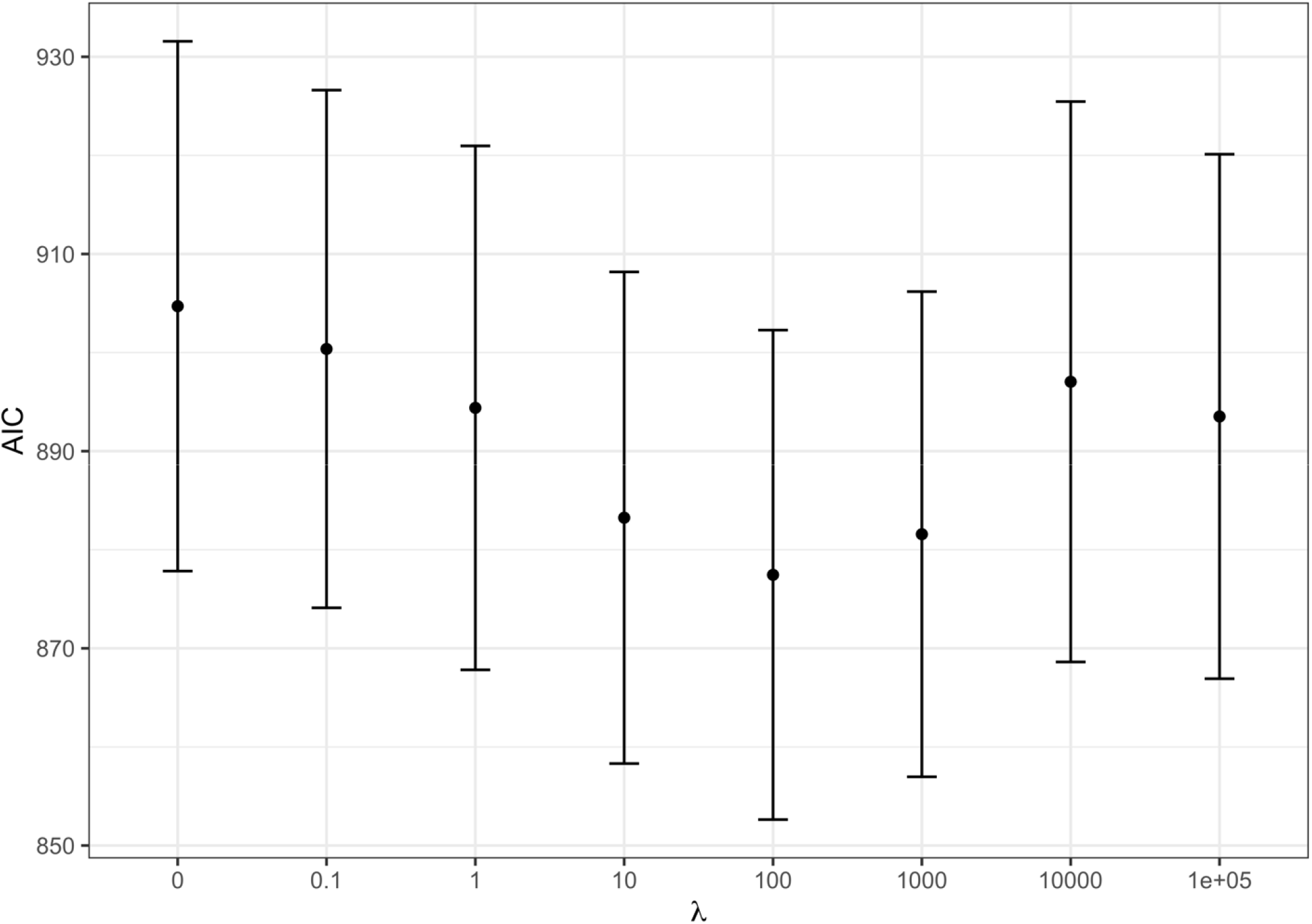
AIC at varying shrinkage parameters: WQS regression Akaike Information Criterion (AIC) at varying shrinkage parameter. The red dots represent the average AIC obtained by fitting WQS models on the 100 datasets fixing to the corresponding value.

We then checked the accuracy of the parameter estimates at varying shrinkage values. In figure 3 the bias of the regression parameters *β*_1*p*_ and *β*_1*n*_ is presented for the positive and the negative direction respectively at varying. The most accurate estimates for the regression parameters correspond to the shrinkage parameter that minimized the AIC (*λ* = 100).

**Figure 3.**
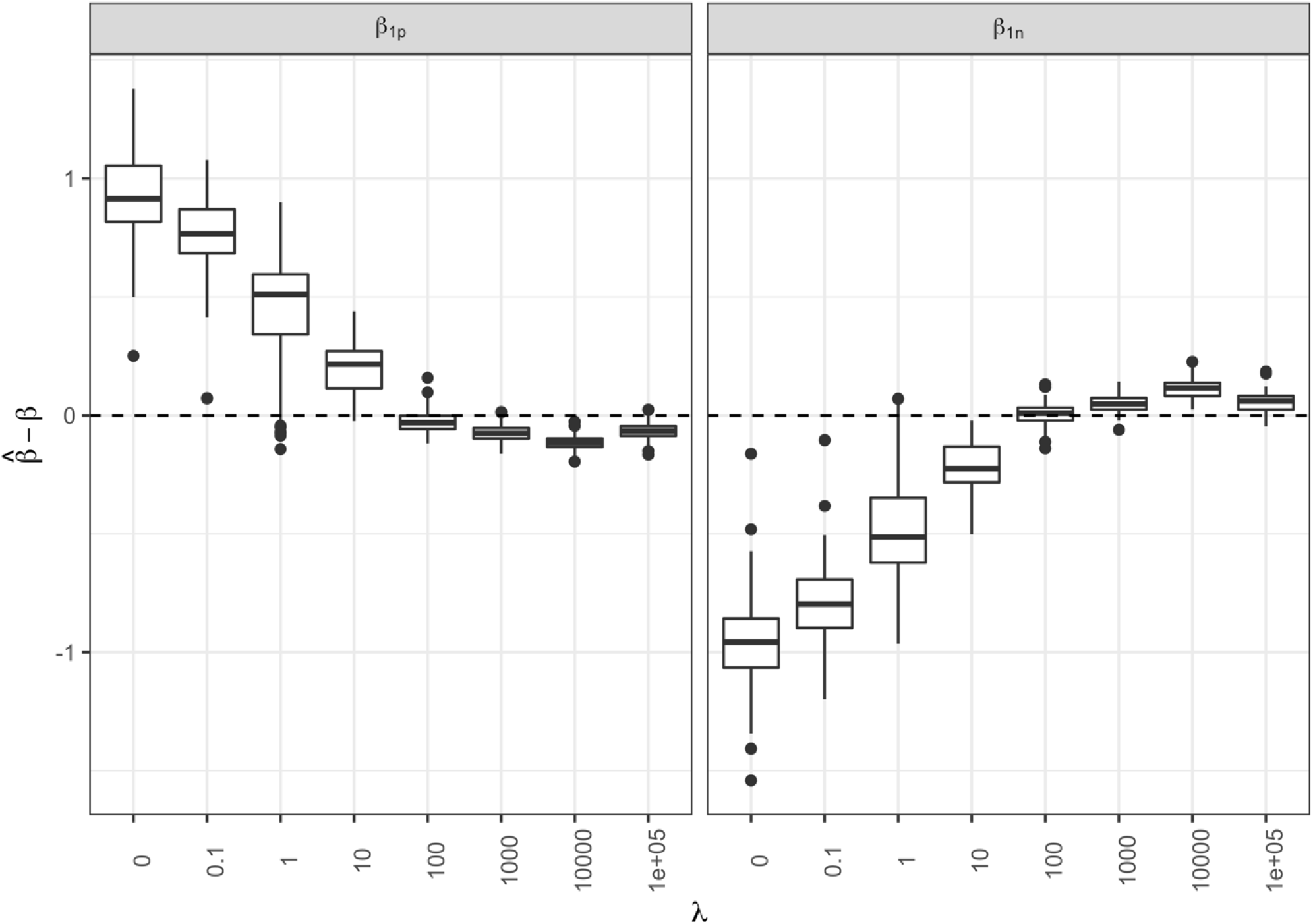
Regression parameter estimates at varying shrinkage parameters: Box-plots of the bias associated to the and regression parameter estimates for the positive and negative direction respectively at different shrinkage parameters

We performed the same analysis for the weights: in figure 4 we can see that when we *λ* = 100 we have accurate estimates both in terms of average sensitivity and specificity in identifying the elements truly associated with the outcome and those that do not have any relationship both for the positive (panel A) and negative (panel B) direction (positive direction: average sensitivity = 49.5%, average specificity = 96.7%; negative direction: average sensitivity = 69.0%, average specificity = 88.3%). The relatively low sensitivity was mainly due to the small number of elements truly associated with the outcome.

**Figure 4.**
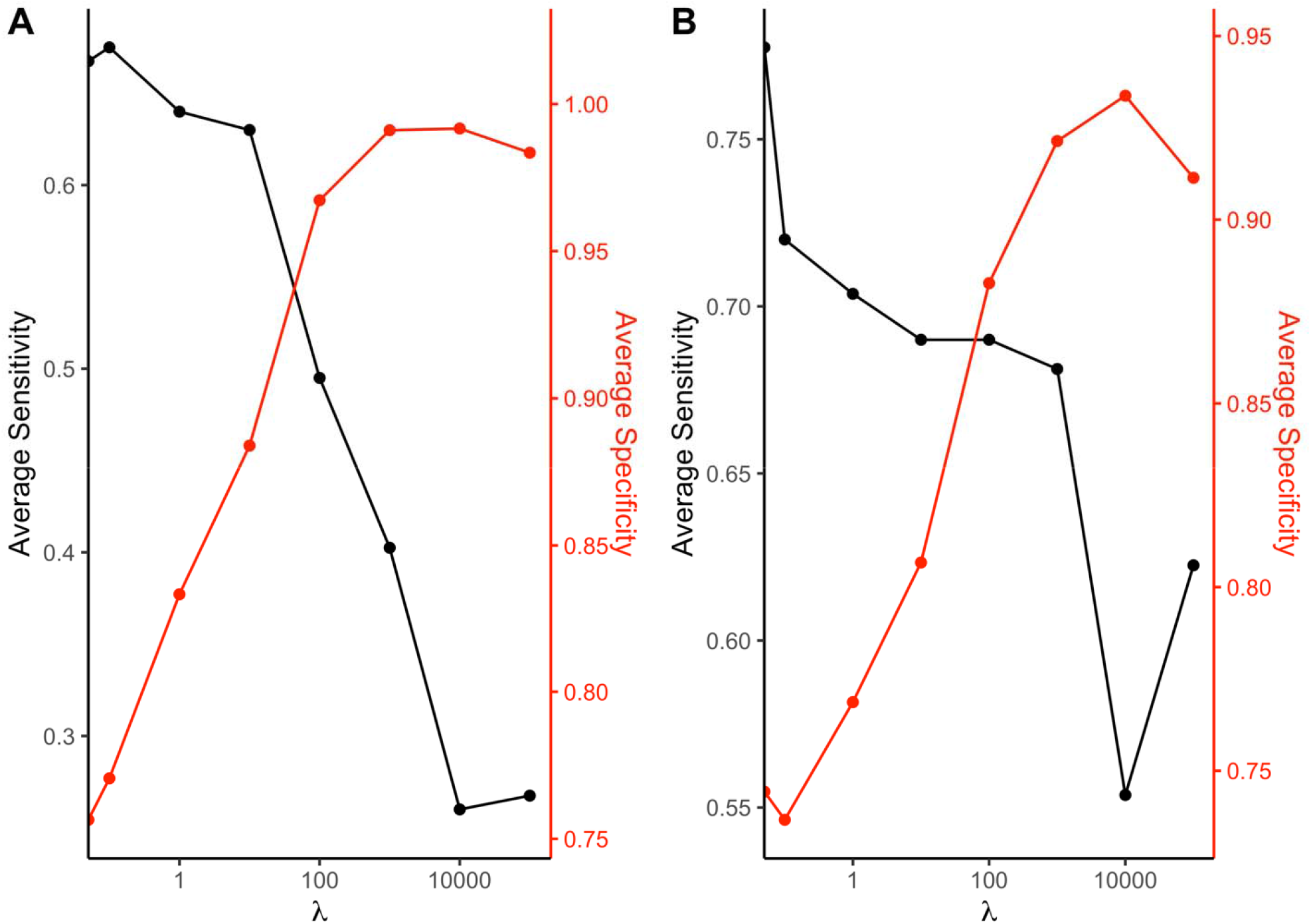
Sensitivity and specificity at varying shrinkage parameter: Average sensitivity and specificity of the 2iWQS method in detecting the elements with a weight greater than 0 and those with null weight associated to a positive (panel A) or a negative (panel B) direction at varying.

Once the optimum shrinkage parameter *λ* was identified, three additional regressions were fitted on each of the 100 datasets: in method 1 two separate WQS regressions were performed, one exploring the positive direction (i.e., where *β*_1_ was constrained to be positive in the nonlinear estimation) and the second exploring the negative direction; in method 2 the WQS set of weights was estimated separately for the positive and negative directions and once the WQS indices were estimated they were included in the same regression model fitted on the validation set. We then compared these results with the ones obtained applying the 2iWQS regression and penalized weights which we will refer to as method 3 from now on.

Figure 5 shows the box-plots of the bias associated with the *β*_1*p*_ and *β*_1*n*_ for method 1, 2 and 3. We can see how model 3 is able to give a better estimate of the parameters (Positive direction Median Error (PME): 0.017, Standard error (SE): 0.055; Negative direction Median Error (NME): -0.023, SD: 0.048) compared to method 1 (PME: -0.141, SE: 0.038; NME: 0.078, SD: 0.047) and method 2 (PME: -0.123, SE: 0.033; NME: 0.054, SD: 0.038).

**Figure 5.**
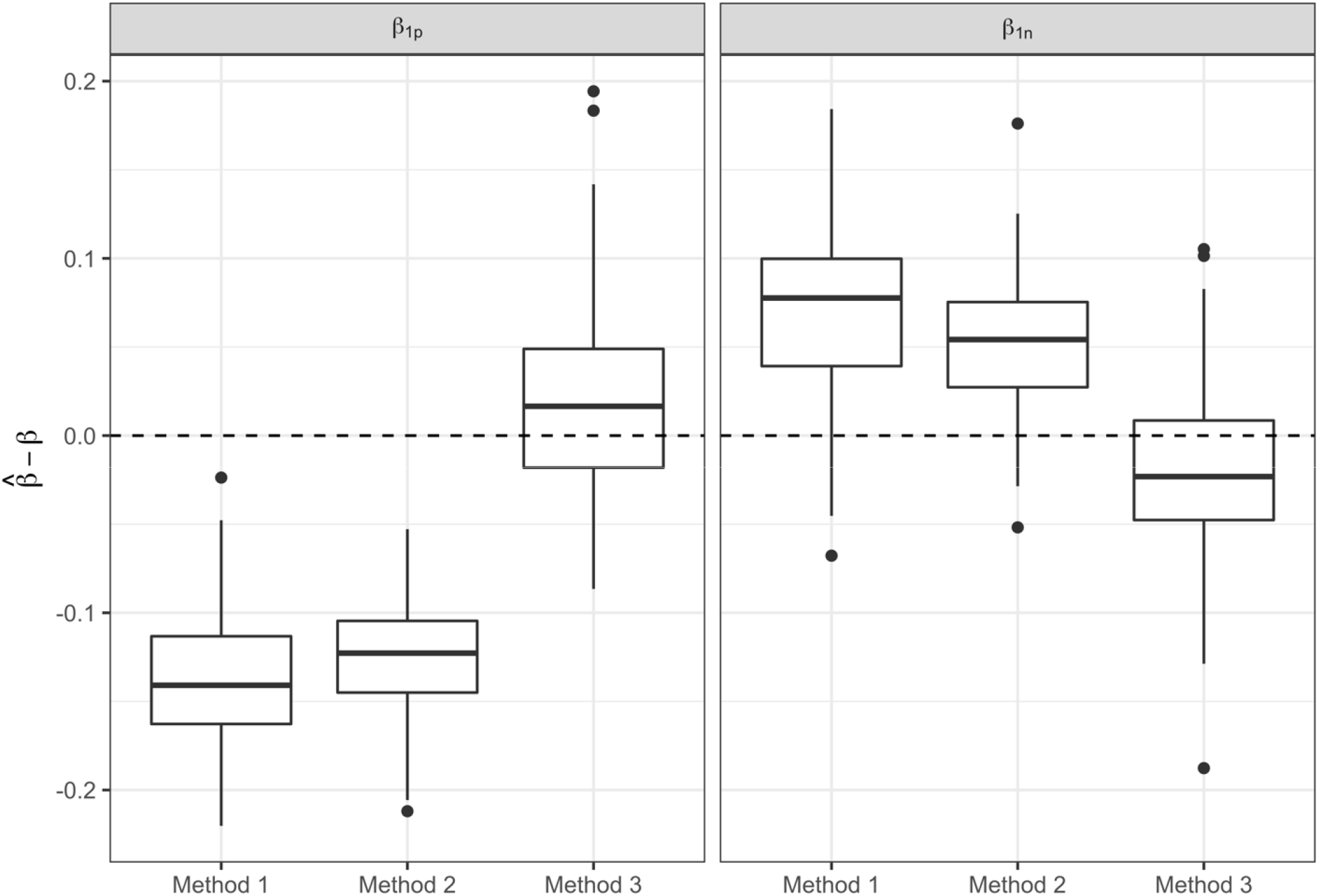
Regression parameter estimates of the three methods: Box-plots of the bias in the regression parameter estimates associated with the two WQS indices of the three methods.

To measure the performance of the 3 methods in identifying the elements associated with the outcome and those that do not have any relationship (in this case method 1 and 2 share the same weight estimates) we considered the average sensitivity and specificity. When we set a threshold equal to the inverse of the number of elements in the mixture to discriminate between the significant and non-significant weights, we observed that method 3 shows better sensitivity in the positive direction compared to methods 1 and 2 (method 1-2 = 26.3%, method 3 = 56.5%) while the sensitivity in the negative direction (method 1-2 = 68.5%, method 3 = 66.3%) and the specificity in both directions were similar (positive direction: method 1-2 = 97.1%, method 3 = 94.7%; negative direction: method 1-2 = 87.4%, method 3 = 87.6%) (figure S1 and S2).

The same analysis was performed in a different scenario where the correlation values among the elements in the mixture were halved. Similar results were obtained in this scenario compared to the one considering the original correlation matrix: low bias was observed in estimating beta regression parameters in the new scenario for method 3 (PME: 0.008, SE: 0.053; NME: 0.015, SE: 0.065), while method 2 showed better estimates (PME: -0.057, SE: 0.045; NME: 0.004, SE: 0.050) (figure S3).

When we looked at the ability of the methods in detecting the true elements with a non-null weight and those not associated with the outcome we observed a better average sensitivity of method 3 compared to method 1-2 (method 1-2 = 42.8%, method 3 = 59.3%) and similar specificity (method 1-2 = 95.5%, method 3 = 96.8%) in the positive direction. In the negative direction method 3 showed a higher specificity (method 1-2 = 86.6%, method 3 = 93.0%) but a lower sensitivity (method 1-2 = 74.1%, method 3 = 66.0%) compared to method 1-2 (figure S4 and S5).

In the last analysis, we tested the performance of the penalized weights when considering a unidirectional association between the mixture and the outcome. In this case, we only applied method 1 and 3 but this last model included a single index and a penalization term was fixed at *λ* = 1000. We note that the 2iWQS regression model cannot be estimated if there is a signal in a single direction. An error message saying that no association is found in one of the two directions is shown by the function. A single index WQS regression in the direction where the signal was detected can be performed in this case. The method with the penalization term showed better estimates of the regression parameter associated with the WQS index (method 3, ME: 0.003, SE: 0.034) (figure S6). Method 3 showed lower sensitivity (method 1 = 63.8%, method 3 = 56.5%) but a higher specificity (method 1 = 90.7%, method 3 = 96.0%) (figure S7).

### Case study

In total 5105 subjects were included in the case study analysis. Table 2 shows the descriptive statistics of the overall population and divided by obese and non-obese for the covariates included in the 2iWQS regression. A total of 1337 (26.2%) subjects were obese and were characterized by a higher median age and higher prevalence of females compared to non-obese participants. A different race distribution was also observed showing a higher percentage of Mexican and Black subjects and a lower prevalence of Asian participants among obese. Finally, a lower level of education was detected among obese people as well as a lower income to poverty ratio index and a higher frequency of low time spent performing moderate to vigorous-intensity activities.

**Table 2.** Sociodemographics and lifestyles: Descriptive statistics of the variables included in the study for the overall population and divided by obese and non-obese. Median, 1^st^ and 3^rd^ quartiles are shown for continuous variables while counts and percentages were considered for categorical variables. Kruskal-Wallis test and Chi-squared test were used to test for differences for continuous and categorical variables respectively.

|  | **Non-Obese (N=3768)** | **Obese (N=1337)** | **Total (N=5105)** | **p value** |
| --- | --- | --- | --- | --- |
| **Age** | 38.0 (28.0, 48.0) | 42.0 (32.0, 51.0) | 39.0 (29.0, 49.0) | < 0.001 |
| **Sex** |  |  |  | 0.010 |
| Males | 1992 (52.9%) | 652 (48.8%) | 2644 (51.8%) |  |
| Females | 1776 (47.1%) | 685 (51.2%) | 2461 (48.2%) |  |
| **Race** |  |  |  | < 0.001 |
| Mexican | 425 (11.3%) | 241 (18.0%) | 666 (13.0%) |  |
| Other Hispanic | 347 (9.2%) | 148 (11.1%) | 495 (9.7%) |  |
| White | 1499 (39.8%) | 501 (37.5%) | 2000 (39.2%) |  |
| Black | 664 (17.6%) | 322 (24.1%) | 986 (19.3%) |  |
| Asian | 695 (18.4%) | 70 (5.2%) | 765 (15.0%) |  |
| Others | 138 (3.7%) | 55 (4.1%) | 193 (3.8%) |  |
| **Education** | 4.0 (3.0, 5.0) | 4.0 (3.0, 4.0) | 4.0 (3.0, 5.0) | < 0.001 |
| **Family income to poverty ratio** | 2.4 (1.1, 4.6) | 2.1 (1.1, 3.8) | 2.3 (1.1, 4.4) | 0.001 |
| **Minutes of sedentary activity** | 360.0 (240.0, 480.0) | 360.0 (240.0, 480.0) | 360.0 (240.0, 480.0) | 0.442 |
| **Moderate and Vigorous-Intensity Activities** |  |  |  | <0.001 |
| Low | 1231 (32.7%) | 517 (38.7%) | 1748 (34.2%) |  |
| Medium | 1319 (35.0%) | 400 (29.9%) | 1719 (33.7%) |  |
| High | 1218 (32.3%) | 420 (31.4%) | 1638 (32.1%) |  |
| **Smoking status** |  |  |  | 0.664 |
| Never | 2301 (61.1%) | 800 (59.8%) | 3101 (60.7%) |  |
| Former | 607 (16.1%) | 228 (17.1%) | 835 (16.4%) |  |
| Current | 860 (22.8%) | 309 (23.1%) | 1169 (22.9%) |  |

In table 3 are shown the summary statistics related to the 38 nutrients included in the analysis. All the elements that showed a significant difference between the two groups had higher values among non-obese subjects.

**Table 3.** Nutrient characteristics: Summary statistics of the 38 nutrients included in the analysis. Median, 1^st^ and 3^rd^ quartiles are shown for the overall population and divided in obese and non-obese participants. Kruskal-Wallis test was applied to test for differences between the

|  | Non-Obese (N=3768) | Obese (N=1337) | Total (N=5105) | p value |
| --- | --- | --- | --- | --- |
| Added vitamin B12 (mcg) | 0.0 (0.0, 1.2) | 0.0 (0.0, 0.9) | 0.0 (0.0, 1.2) | 0.063 |
| Alcohol (gm) | 0.0 (0.0, 7.8) | 0.0 (0.0, 0.0) | 0.0 (0.0, 7.0) | < **0.001** |
| Alpha-carotene (mcg) | 69.8 (20.0, 442.0) | 56.0 (17.0, 293.0) | 65.5 (19.5, 408.0) | < **0.001** |
| Beta-carotene (mcg) | 1090.5 (417.2, 2978.0) | 838.0 (348.5, 2066.0) | 1024.0 (396.0, 2711.5) | < **0.001** |
| Caffeine (mg) | 94.0 (25.5, 191.0) | 102.5 (26.0, 209.0) | 95.5 (26.0, 193.0) | 0.056 |
| Calcium (mg) | 958.5 (656.0, 1358.0) | 940.5 (641.5, 1309.0) | 954.0 (652.5, 1348.7) | 0.140 |
| Carbohydrate (gm) | 254.6 (190.3, 333.0) | 245.1 (179.9, 320.2) | 251.1 (188.1, 330.0) | **0.002** |
| Cholesterol (mg) | 255.5 (158.0, 394.5) | 254.0 (163.5, 416.0) | 254.5 (160.0, 399.0) | 0.167 |
| Choline (mg) | 314.1 (224.1, 435.0) | 305.2 (210.7, 434.4) | 312.4 (220.9, 434.6) | **0.045** |
| Copper (mg) | 1.3 (0.9, 1.8) | 1.2 (0.8, 1.7) | 1.2 (0.9, 1.8) | < **0.001** |
| Beta-cryptoxanthin (mcg) | 38.5 (12.5, 101.1) | 41.5 (12.5, 99.5) | 39.0 (12.5, 100.0) | 0.678 |
| Dietary fiber (gm) | 16.4 (11.2, 23.8) | 15.2 (10.2, 22.0) | 16.1 (10.9, 23.3) | < **0.001** |
| Folate, DFE (mcg) | 606.2 (403.9, 948.8) | 529.0 (350.5, 838.5) | 589.0 (388.5, 921.5) | < **0.001** |
| Iron (mg) | 15.1 (10.9, 21.7) | 14.1 (10.0, 20.5) | 14.8 (10.7, 21.3) | < **0.001** |
| Lutein + zeaxanthin (mcg) | 890.0 (472.8, 1819.6) | 811.5 (442.5, 1537.0) | 869.0 (464.0, 1735.0) | < **0.001** |
| Lycopene (mcg) | 2536.5 (647.8, 6617.9) | 2514.0 (694.0, 7188.5) | 2528.5 (657.0, 6829.5) | 0.665 |
| Magnesium (mg) | 307.0 (229.5, 413.6) | 285.0 (206.0, 379.0) | 301.0 (222.0, 406.0) | < **0.001** |
| Monounsaturated fatty acids (gm) | 26.9 (18.8, 36.5) | 26.5 (19.0, 36.1) | 26.8 (18.8, 36.4) | 0.760 |
| Niacin (mg) | 28.5 (20.2, 40.4) | 26.8 (19.3, 38.0) | 28.1 (19.9, 39.6) | < **0.001** |
| Phosphorus (mg) | 1352.2 (1023.5, 1759.5) | 1291.0 (991.0, 1724.0) | 1338.0 (1016.5, 1750.5) | **0.018** |
| Polyunsaturated fatty acids (gm) | 17.4 (11.9, 24.3) | 17.5 (12.2, 24.7) | 17.4 (12.0, 24.4) | 0.576 |
| Potassium (mg) | 2600.8 (1955.8, 3361.6) | 2418.5 (1837.0, 3160.5) | 2563.2 (1921.5, 3327.0) | < **0.001** |
| Protein (gm) | 81.7 (61.4, 107.5) | 78.1 (58.3, 103.1) | 80.9 (60.5, 106.6) | **0.005** |
| Retinol (mcg) | 2.2 (1.5, 3.3) | 2.1 (1.4, 3.2) | 2.2 (1.5, 3.3) | **0.002** |
| Riboflavin (Vitamin B2) (mg) | 24.1 (16.2, 34.1) | 24.1 (16.9, 34.0) | 24.1 (16.3, 34.1) | 0.545 |
| Saturated fatty acids (gm) | 122.9 (89.1, 166.2) | 118.0 (85.1, 160.0) | 121.6 (88.0, 164.8) | **0.013** |
| Selenium (mcg) | 3480.8 (2582.4, 4578.6) | 3396.0 (2583.0, 4433.0) | 3468.0 (2582.5, 4549.5) | 0.172 |
| Sodium (mg) | 100.4 (65.8, 145.3) | 99.0 (63.7, 146.7) | 100.0 (65.1, 145.9) | 0.591 |
| Sugars (gm) | 7.5 (0.0, 41.5) | 6.0 (0.0, 40.0) | 7.0 (0.0, 41.0) | 0.056 |
| Theobromine (mg) | 1.8 (1.3, 2.7) | 1.7 (1.2, 2.5) | 1.8 (1.2, 2.6) | < **0.001** |
| Thiamin (Vitamin B1) (mg) | 516.0 (304.5, 815.5) | 457.5 (274.0, 730.0) | 502.5 (296.0, 788.0) | < **0.001** |
| Vitamin B12 (mcg) | 5.5 (3.1, 10.6) | 5.1 (2.8, 9.7) | 5.4 (3.0, 10.4) | **0.008** |
| Vitamin B6 (mg) | 2.4 (1.6, 3.9) | 2.1 (1.4, 3.4) | 2.3 (1.5, 3.8) | < **0.001** |
| Vitamin C (mg) | 86.5 (37.5, 169.5) | 74.8 (30.6, 147.0) | 83.8 (35.6, 161.5) | < **0.001** |
| Vitamin D (D2 + D3) (mcg) | 5.6 (2.3, 13.7) | 4.6 (2.1, 11.3) | 5.3 (2.2, 13.2) | < **0.001** |
| Vitamin E as alpha-tocopherol (mg) | 7.7 (5.3, 11.3) | 7.4 (5.0, 10.8) | 7.7 (5.3, 11.2) | **0.007** |
| **Vitamin K (mcg)** | 86.9 (51.4, 153.9) | 76.6 (45.8, 128.5) | 84.3 (49.8, 147.1) | **< 0.001** |
| **Zinc (mg)** | 12.3 (8.4, 18.3) | 11.5 (7.7, 17.2) | 12.1 (8.2, 18.0) | **< 0.001** |

We then applied the 2iWQS regression to test for the association between the nutrients and the outcome adjusting for all the covariates reported in Table 2. We used a repeated holdout approach to have more stable results including all the observations in the study to estimate both the weights and the regression parameters during the repeated testing and validation steps (Tanner et al., 2019). A total of 100 repeated holdout 2iWQS regressions were performed. Table 4 shows the effects and their 95% Confidence Intervals (CIs) of both the positive and the negative index on the probability of being obese. Both indices were associated with the outcome. The median was used as the parameter point estimates while the 2.5^th^ and the 97.5^th^ percentiles were considered to build the 95% CIs. In table 4 are also shown the medians of the weights greater than the prespecified cutoff (1/38=0.026) for the positive and negative index where caffeine and magnesium showed a predominant role in the positive and negative association with obesity respectively. Moreover, these two nutrients showed no weights below the threshold set to 0.026. The distributions of all the elements included in the analysis estimated for both indices are provided in Figure 6.

**Figure 6.**
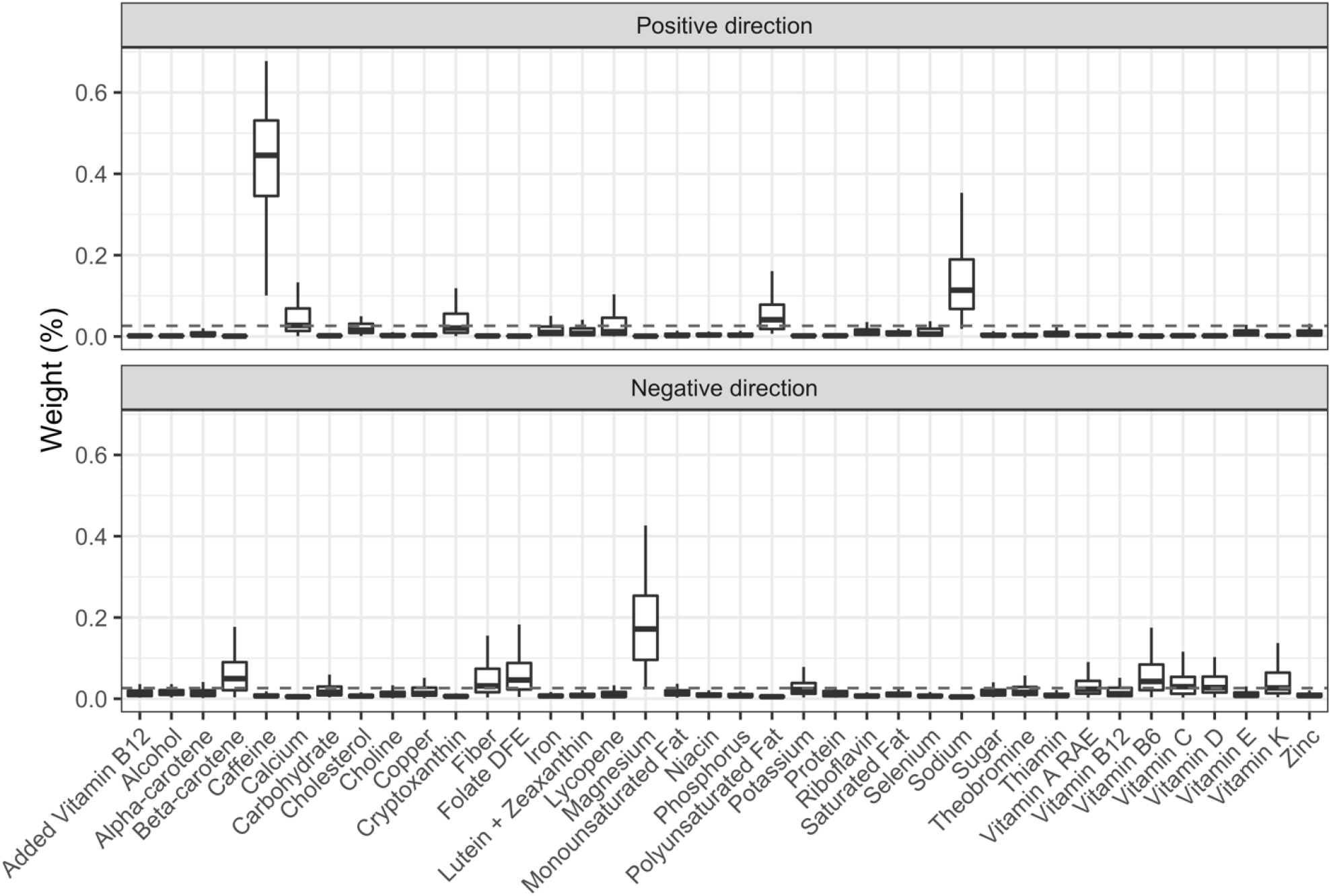
Sensitivity and specificity of the three methods: Box-plot of the weights associated to the positive and negative index estimate through the repeated holdout 2iWQS regression. The dashed line represents the prespecified cut-off established to identify the most important elements of the mixture and set equal to the inverse of the number of the mixture components (0.026).

**Table 4.** 2iWQS regression results: The estimates and 95% confidence intervals (CI) of the positive (PWQS) and negative (NWQS) indices are shown. Model was adjusted for age, sex, race, education, the ratio of family income to poverty, the minutes of sedentary activity, the minutes of moderate and vigorous activities and the smoking status. The second part of the table shows the magnitude of the weights greater than the prespecified cutoff (0.026) for both the positive and the negative index.

|  | **Estimate** | **95% CI** |
| --- | --- | --- |
| PWQS | 0.069 | 0.023; 0.153 |
| NWQS | -0.114 | -0.176; -0.066 |
| **Weights** |  |  |
| **PWQS** |  |  |
| Caffeine | 0.445 |  |
| Sodium | 0.114 |  |
| Polyunsaturated Fat | 0.041 |  |
| Calcium | 0.027 |  |
| **NWQS** |  |  |
| Magnesium | 0.172 |  |
| Beta-carotene | 0.050 |  |
| Folate DFE | 0.046 |  |
| Vitamin B6 | 0.043 |  |
| Fiber | 0.032 |  |
| Vitamin C | 0.030 |  |
| Vitamin D | 0.028 |  |
| Vitamin K | 0.027 |  |

Abbreviations: CI, Confidence Interval; DFE, Dietary Folate Equivalents; PWQS, Positive
Weighted Quantile Sum index; NWQS, Negative Weighted Quantile Sum index.

## Discussion

Through this work we were able to extend the WQS regression to the case where the mixture considered in the study can have both a positive and a negative effect, moreover, we increased the ability of WQS in detecting the association between the mixture and the outcome as well as in identifying the true elements through the introduction of penalized weight estimates. The new method estimates two indices in the same regression model both including all the elements of the mixture, one constrained to be positive and one to be negative. A recent work from Keil and others(2020) (Keil et al., 2020) introduced a new approach that estimates the overall effect of the mixture on the outcome when there is uncertainty about the effect direction of some exposures. Differently from the original WQS regression, they proposed to estimate positive and negative weights within the same index using normalized linear (or generalized linear) regression coefficients and then estimating the mixture effect via a standard g-computation algorithm. With this approach, we are able to measure the overall effect of the mixture (which is not the mixture effect) on the outcome but we cannot estimate the impact in the positive and negative directions. Through our method, we introduced the possibility to measure both the beneficial and harmful effects of the exposure to the mixture separately keeping the advantages of the identification of a weighted index that allows increasing the power to detect the mixture effect and avoiding incurring the reversal paradox. Another advantage that we carry from WQS regression compared to other shrinkage methods is the ability to estimate an overall mixture effect, in our case in both directions, and to avoid in presence of highly correlated variables a grouping effect like in elastic net or the arbitrary selection of variables like in LASSO that can be problematic in the risk evaluation of environmental mixture (Carrico et al., 2015).

In this work, we showed how the two indices were built and how we deal with the correlation between the two indices. This was the main issue that we encountered: because of the high correlation among the elements included in the mixture we noticed a risk of collinearity when including both indices in the same regression model. To face this problem, we applied two strategies. As a first solution, we introduced a penalization parameter in the objective function to estimate the weights allowing us to better discriminate between the elements that have a weight significantly different from zero and those that have a null weight. This helps to reduce the noise produced by the elements that are not associated with the outcome which can increase the correlation between the two indices. The second solution to reduce the correlation between the two indices was the application of the weighted mean based on the tolerance of each bootstrapped model in the estimates of the final weights. This gave more importance to the set of weights that produces less correlated indices.

In the real case study, we applied this new methodology by taking data from the NHANES 2011-2012, 2013-2014 and 2015-2016 study cycles to test for the association between nutrients and obesity. The results showed a harmful effect of caffeine, calcium, polyunsaturated fatty acids and sodium. While some nutrients like sodium (Kang et al., 2016; Lee et al., 2018; Zhou et al., 2019) are already known risk elements for obesity, we also found nutrients for which there is controversial evidence of their effect on obesity. Because of the unavailable information on the different types of polyunsaturated fatty acids we were not able to disentangle which component drove the harmful effect on obesity. Polyunsaturated fatty acids are known to be protective against overweight and obesity (Albracht-Schulte et al., 2018; Ralston et al., 2017; Rogero and Calder, 2018; Saini and Keum, 2018; Silva Figueiredo et al., 2017; Tortosa-Caparrós et al., 2017), however, there is evidence that an increased intake of omega-6 long-chain polyunsaturated fatty acids can increase the risk of obesity (Fekete et al., 2015; Saini and Keum, 2018) in particular if there is an unbalanced omega-6/omega-3 ratio, an increasingly widespread problem in western countries (Simopoulos, 2016). Calcium intake was observed to be a protective factor against obesity (de Oliveira Freitas et al., 2012; Pannu et al., 2016; Villarroel et al., 2014; Zhang et al., 2019) but few studies did not show any effect (Soares et al., 2011; Song and Sergeev, 2012) or a harmful relationship (Sadeghi et al., 2018). Finally, caffeine has a protective effect on a regular intake (Bhatti et al., 2013) but in excessive doses, it can affect insomnia and anxiety (Nehlig, 2018; Yang et al., 2010) which are associated in turn with obesity (Amiri and Behnezhad, 2019; Cai et al., 2018; Rajan and Menon, 2017). On the other hand, a protective effect against obesity was found for magnesium, beta-carotene, folate Dietary Folate Equivalents (DFE), vitamin B6, fiber, vitamin C, vitamin D and vitamin K. For all of these nutrients there was evidence of a beneficial effect against obesity in previous studies (Al-Suhaimi and Al-Jafary, 2020; Astrup and Bügel, 2019; Bonet et al., 2016; Cho et al., 2013; Coronel et al., 2019; Garcia-Diaz et al., 2014; Oliveira et al., 2017; Pereira-Santos et al., 2015; Perveen et al., 2015; Piuri et al., 2021; Pourshahidi, 2015; Savastano et al., 2017; Thomas-Valdés et al., 2017; Thompson et al., 2017; Walsh et al., 2017; □erban et al., 2019).

One strength of this novel approach is the ability to include all nutrients in the analysis considering the possible confounding that can be caused by the exclusion of some elements while previous studies showed the association of one or few elements at a time with obesity. Moreover, we showed that considering two indices in the same WQS regression with the penalization term increased the accuracy of the parameter estimates when the mixture has a bidirectional effect on the outcome of interest. In addition to already available methods like the quantile-based g-computation approach, our new methodology allows us to measure the double association of the mixture quantifying the effect in both the positive and negative direction with the dependent variable. As for WQS regression, a further advantage of this approach is the ease of use and of interpretation of the results due to the building of the indices representing the mixture exposure and the weights attributed to each element identifying the contribution of each element to the estimated association with the outcome. One additional advantage to be considered is the possibility to integrate this new feature of WQS in the random subset (Curtin et al., 2019) and repeated holdout (Tanner et al., 2019) extensions.

In contrast to the simplicity of the method, we can identify as a limitation of the WQS regression the lower flexibility due to the assumption of a linear trend between each element and the dependent variable. Before applying this method, it is recommended to check in advance if a non-linear trend exists between the mixture components and the outcome by applying other regression methods that can deal with environmental mixtures like Bayesian Kernel Machine Regression (Bobb et al., 2015).

## Conclusion

Through this work, we introduced an extension of the WQS regression that showed the possibility to improve the accuracy of the parameter estimates in WQS with a single index as well as when considering a mixture of elements that can have both a protective and a harmful effect on the outcome. This was possible through the application of penalized weight estimates and the introduction of a second index, allowing one WQS index to investigate the positive and one the negative direction of the association with the dependent variable. The usage of two indices in the same model enables to quantify the effect of the mixture in the two directions keeping them separate. This can be of great interest in fields like nutrition where a measure of the protective and harmful effects of nutrients can help to take decisions on the diet of patients.

## Data Availability

All data in the present study are available within the gWQS R package (Renzetti et al.).

https://github.com/renzetti/g2iWQS/blob/main/data/tiwqs_data.RData

## Abbreviations

2iWQS: two-indices WQS
AIC: Akaike Information Criterion
BMI: Body Mass Index
CI: Confidence Interval
DFE: Dietary Folate Equivalents
NHANES: National Health and Nutrition Examination Survey
NME: Negative direction Median Error
PME: Positive direction Median Error
SE: Standard Error
VIF: Variance Inflation Factor
WQS: Weighted Quantile Sum

## Acknowledgments

SC work was supported by research grants from the Italian Ministry of University (PRIN projects n. 20178S4EK9). CG was supported by the National Institute of Environmental Health Sciences (#U2CES026555 and #R01ES028811)

## Supplementary materials, code and research data

The reader is referred to the on-line Supplementary Materials for additional results and R code. Data are available within the gWQS R package (Renzetti et al.).

## Notes

Conflict of interest *The authors declare they have nothing to disclose.*

### Competing Interest Statement

The authors have declared no competing interest.

### Author Declarations

The study used ONLY simulated and openly available human data that were originally located at: https://wwwn.cdc.gov/nchs/nhanes/

## References

National Health and Nutrition Examination Survey (NHANES). Phone Follow-Up Dietary Interviewer Procedures Manual 2016a.

National Health and Nutrition Examination Survey (NHANES). MEC In-Person Dietary Interviewers Procedures Manual., 2016b.

Al-Suhaimi, E. A., Al-Jafary, M. A., 2020. Endocrine roles of vitamin K-dependent-osteocalcin in the relation between bone metabolism and metabolic disorders. Rev Endocr Metab Disord. 21, 117–125.

Albracht-Schulte, K., et al., 2018. Omega-3 fatty acids in obesity and metabolic syndrome: a mechanistic update. J Nutr Biochem. 58, 1–16.

Amiri, S., Behnezhad, S., 2019. Obesity and anxiety symptoms: a systematic review and meta-analysis. Neuropsychiatr. 33, 72–89.

Astrup, A., Bügel, S., 2019. Overfed but undernourished: recognizing nutritional inadequacies/deficiencies in patients with overweight or obesity. Int J Obes (Lond). 43, 219–232.

Bhatti, S. K., et al., 2013. Coffee and tea: perks for health and longevity? Curr Opin Clin Nutr Metab Care. 16, 688–97.

Billionnet, C., et al., 2012. Estimating the health effects of exposure to multi-pollutant mixture. Ann Epidemiol. 22, 126–41.

Bobb, J. F., et al., 2015. Bayesian kernel machine regression for estimating the health effects of multi-pollutant mixtures. Biostatistics. 16, 493–508.

Bonet, M. L., et al., 2016. Carotenoids in Adipose Tissue Biology and Obesity. Subcell Biochem. 79, 377–414.

Braun, J. M., et al., 2016. What Can Epidemiological Studies Tell Us about the Impact of Chemical Mixtures on Human Health? Environ Health Perspect. 124, A6–9.

Bretz, F., et al., 2005. Combining multiple comparisons and modeling techniques in dose-response studies. Biometrics. 61, 738–48.

Cai, G. H., et al., 2018. Insomnia symptoms and sleep duration and their combined effects in relation to associations with obesity and central obesity. Sleep Med. 46, 81–87.

Carlin, D. J., et al., 2013. Unraveling the health effects of environmental mixtures: an NIEHS priority. Environ Health Perspect. 121, A6–8.

Carpenter, D. O., et al., 2002. Understanding the human health effects of chemical mixtures. Environ Health Perspect. 110 Suppl 1, 25–42.

Carrico, C., et al., 2015. Characterization of Weighted Quantile Sum Regression for Highly Correlated Data in a Risk Analysis Setting. J. Agric. Biol. Environ. Stat. 20, 100–120.

Cho, S. S., et al., 2013. Consumption of cereal fiber, mixtures of whole grains and bran, and whole grains and risk reduction in type 2 diabetes, obesity, and cardiovascular disease. Am J Clin Nutr. 98, 594–619.

Coronel, J., et al., 2019. β-carotene in Obesity Research: Technical Considerations and Current Status of the Field. Nutrients. 11.

Curtin, P., et al., A random subset implementation of weighted quantile sum (WQSRS) regression for analysis of high-dimensional mixtures. Vol. 50, 2019, pp. 119-1134.

Czarnota, J., et al., 2015. Assessment of weighted quantile sum regression for modeling chemical mixtures and cancer risk. Cancer Inform. 14, 159–71.

de Oliveira Freitas, D. M., et al., 2012. Calcium ingestion and obesity control. Nutr Hosp. 27, 1758–71.

Dominici, F., et al., 2010. Protecting human health from air pollution: shifting from a single-pollutant to a multipollutant approach. Epidemiology. 21, 187–94.

Dormann, C. F., et al., 2013. Collinearity: a review of methods to deal with it and a simulation study evaluating their performance. Ecography. 36, 27–46.

Fekete, K., et al., 2015. Long-chain polyunsaturated fatty acid status in obesity: a systematic review and meta-analysis. Obes Rev. 16, 488–97.

Garcia-Diaz, D. F., et al., 2014. Vitamin C in the treatment and/or prevention of obesity. J Nutr Sci Vitaminol (Tokyo). 60, 367–79.

Greenland, S., 2008. Multiple comparisons and association selection in general epidemiology. Int J Epidemiol. 37, 430–4.

Jain, P., et al., 2018. A multivariate approach to investigate the combined biological effects of multiple exposures. J Epidemiol Community Health. 72, 564–571.

Kang, Y. J., et al., 2016. Associations of Obesity and Dyslipidemia with Intake of Sodium, Fat, and Sugar among Koreans: a Qualitative Systematic Review. Clin Nutr Res. 5, 290–304.

Keil, A. P., et al., 2020. A Quantile-Based g-Computation Approach to Addressing the Effects of Exposure Mixtures. Environ Health Perspect. 128, 47004.

Leal, C., et al., 2012. Multicollinearity in associations between multiple environmental features and body weight and abdominal fat: using matching techniques to assess whether the associations are separable. Am J Epidemiol. 175, 1152–62.

Lee, J., et al., 2018. Associations of urinary sodium levels with overweight and central obesity in a population with a sodium intake. BMC Nutr. 4, 47.

Nehlig, A., 2018. Interindividual Differences in Caffeine Metabolism and Factors Driving Caffeine Consumption. Pharmacol Rev. 70, 384–411.

Oliveira, A. R., et al., 2017. Hypomagnesemia and its relation with chronic low-grade inflammation in obesity. Rev Assoc Med Bras (1992). 63, 156–163.

Pannu, P. K., et al., 2016. Calcium and Vitamin D in Obesity and Related Chronic Disease. Adv Food Nutr Res. 77, 57–100.

Patel, C. J., 2017. Analytic Complexity and Challenges in Identifying Mixtures of Exposures Associated with Phenotypes in the Exposome Era. Curr Epidemiol Rep. 4, 22–30.

Pereira-Santos, M., et al., 2015. Obesity and vitamin D deficiency: a systematic review and meta-analysis. Obes Rev. 16, 341–9.

Perveen, R., et al., 2015. Tomato (Solanum lycopersicum) Carotenoids and Lycopenes Chemistry; Metabolism, Absorption, Nutrition, and Allied Health Claims--A Comprehensive Review. Crit Rev Food Sci Nutr. 55, 919–29.

Piuri, G., et al., 2021. Magnesium in Obesity, Metabolic Syndrome, and Type 2 Diabetes. Nutrients. 13.

Pourshahidi, L. K., 2015. Vitamin D and obesity: current perspectives and future directions. Proc Nutr Soc. 74, 115–24.

Rajan, T. M., Menon, V., 2017. Psychiatric disorders and obesity: A review of association studies. J Postgrad Med. 63, 182–190.

Ralston, J. C., et al., 2017. Fatty Acids and NLRP3 Inflammasome-Mediated Inflammation in Metabolic Tissues. Annu Rev Nutr. 37, 77–102.

Renzetti, S., et al., gWQS: Generalized Weighted Quantile Sum Regression.

Rogero, M. M., Calder, P. C., 2018. Obesity, Inflammation, Toll-Like Receptor 4 and Fatty Acids. Nutrients. 10.

Sadeghi, O., et al., 2018. Association between dairy consumption, dietary calcium intake and general and abdominal obesity among Iranian adults. Diabetes Metab Syndr. 12, 769–775.

Saini, R. K., Keum, Y. S., 2018. Omega-3 and omega-6 polyunsaturated fatty acids: Dietary sources, metabolism, and significance - A review. Life Sci. 203, 255–267.

Savastano, S., et al., 2017. Low vitamin D status and obesity: Role of nutritionist. Rev Endocr Metab Disord. 18, 215–225.

Savitz, D. A., Olshan, A. F., 1995. Multiple comparisons and related issues in the interpretation of epidemiologic data. Am J Epidemiol. 142, 904–8.

Silva Figueiredo, P., et al., 2017. Fatty Acids Consumption: The Role Metabolic Aspects Involved in Obesity and Its Associated Disorders. Nutrients. 9.

Simopoulos, A. P., 2016. An Increase in the Omega-6/Omega-3 Fatty Acid Ratio Increases the Risk for Obesity. Nutrients. 8, 128.

Soares, M. J., et al., 2011. Calcium and vitamin D for obesity: a review of randomized controlled trials. Eur J Clin Nutr. 65, 994–1004.

Song, Q., Sergeev, I. N., 2012. Calcium and vitamin D in obesity. Nutr Res Rev. 25, 130–41.

Stacey, A. W., Czyz, C. N., 2012. Author response: Analysis of the use of multiple comparison corrections in ophthalmology research. Invest Ophthalmol Vis Sci. 53, 5955.

Stafoggia, M., et al., 2017. Statistical Approaches to Address Multi-Pollutant Mixtures and Multiple Exposures: the State of the Science. Curr Environ Health Rep. 4, 481–490.

Sun, Z., et al., 2013. Statistical strategies for constructing health risk models with multiple pollutants and their interactions: possible choices and comparisons. Environ Health. 12, 85.

Tanner, E. M., et al., 2019. Repeated holdout validation for weighted quantile sum regression. MethodsX. 6, 2855–2860.

Thomas-Valdés, S., et al., 2017. Association between vitamin deficiency and metabolic disorders related to obesity. Crit Rev Food Sci Nutr. 57, 3332–3343.

Thompson, S. V., et al., 2017. Effects of isolated soluble fiber supplementation on body weight, glycemia, and insulinemia in adults with overweight and obesity: a systematic review and meta-analysis of randomized controlled trials. Am J Clin Nutr. 106, 1514–1528.

Tortosa-Caparrós, E., et al., 2017. Anti-inflammatory effects of omega 3 and omega 6 polyunsaturated fatty acids in cardiovascular disease and metabolic syndrome. Crit Rev Food Sci Nutr. 57, 3421–3429.

Tu, Y., et al., 2008. Simpson’s Paradox, Lord’s Paradox, and Suppression Effects are the same phenomenon – the reversal paradox. Emerging Themes in Epidemiology. 5.

Vatcheva, K. P., et al., 2016. Multicollinearity in Regression Analyses Conducted in Epidemiologic Studies. Epidemiology (Sunnyvale). 6.

Villarroel, P., et al., 2014. Calcium, obesity, and the role of the calcium-sensing receptor. Nutr Rev. 72, 627–37.

Walsh, J. S., et al., 2017. Vitamin D in obesity. Curr Opin Endocrinol Diabetes Obes. 24, 389–394.

Weisskopf, M. G., et al., 2018. Bias Amplification in Epidemiologic Analysis of Exposure to Mixtures. Environ Health Perspect. 126, 047003.

Yang, A., et al., 2010. Genetics of caffeine consumption and responses to caffeine. Psychopharmacology (Berl). 211, 245–57.

Zhang, F., et al., 2019. Anti-Obesity Effects of Dietary Calcium: The Evidence and Possible Mechanisms. Int J Mol Sci. 20.

Zhou, L., et al., 2019. Salt intake and prevalence of overweight/obesity in Japan, China, the United Kingdom, and the United States: the INTERMAP Study. Am J Clin Nutr. 110, 34–40.

Șerban, C. L., et al., 2019. Assessment of Nutritional Intakes in Individuals with Obesity under Medical Supervision. A Cross-Sectional Study. Int J Environ Res Public Health. 16.

