## Supplemental material for "A weighted quantile sum regression with penalized weights and two indices"

**Figure S1**: Heatmap of the sensitivity of the three methods in detecting the elements with a weight greater than 0 associated to a positive (panel A) or a negative (panel B) direction.

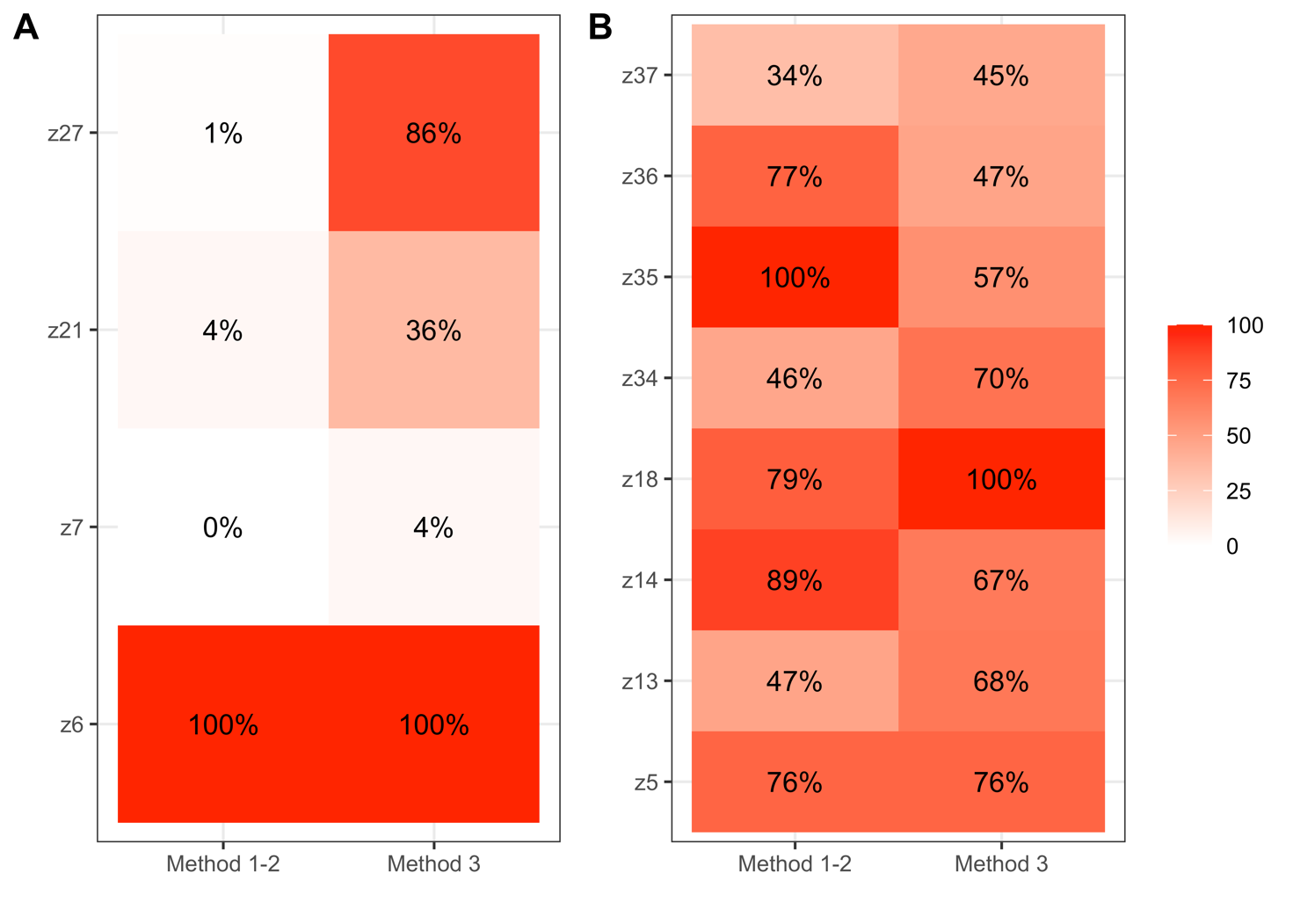

**Figure S2**: Heatmap of the specificity of the three methods in detecting the elements with a null weight associated to a positive (panel A) or a negative (panel B) direction.

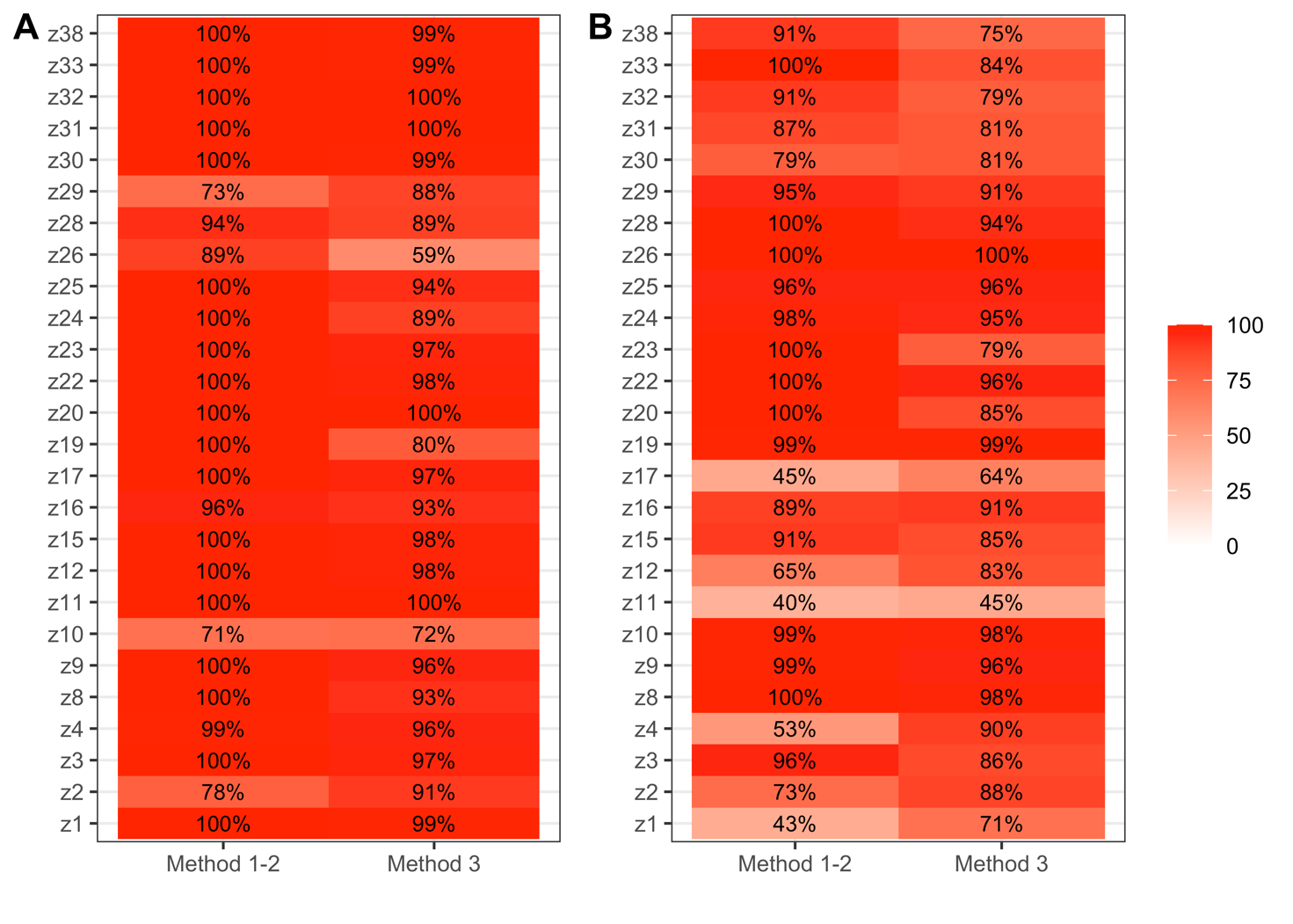

**Figure S3**: Box-plots of the bias in the estimates of the regression parameter associated to the two WQS indices of the three methods in scenario 2 where the correlation values were halved.

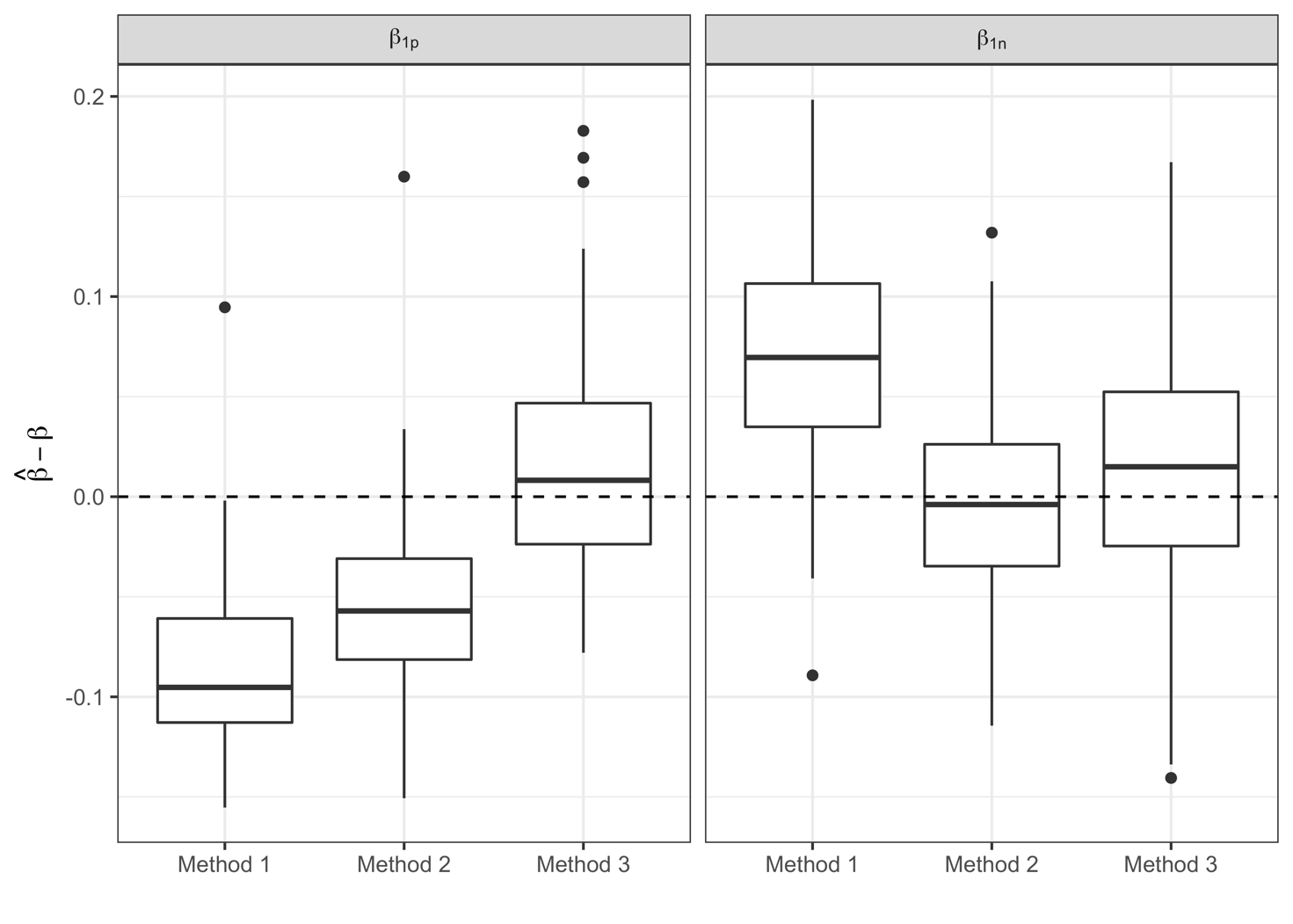

**Figure S4**: Heatmap of the sensitivity of the three methods in detecting the elements with a weight greater than 0 associated to a positive (panel A) or a negative (panel B) direction in scenario 2.

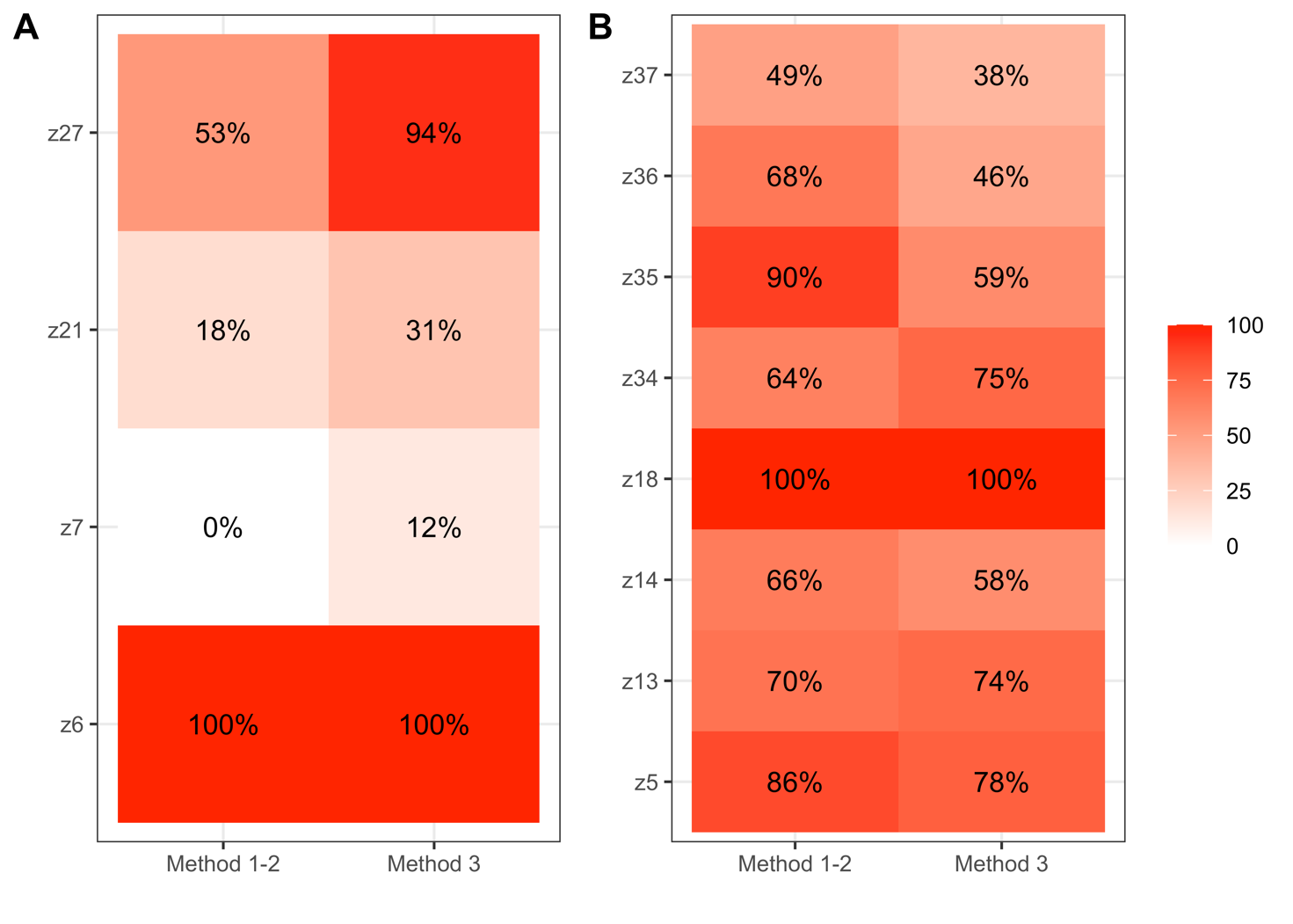

**Figure S5**: Heatmap of the specificity of the three methods in detecting the elements with a null weight associated to a positive (panel A) or a negative (panel B) direction in scenario 2.

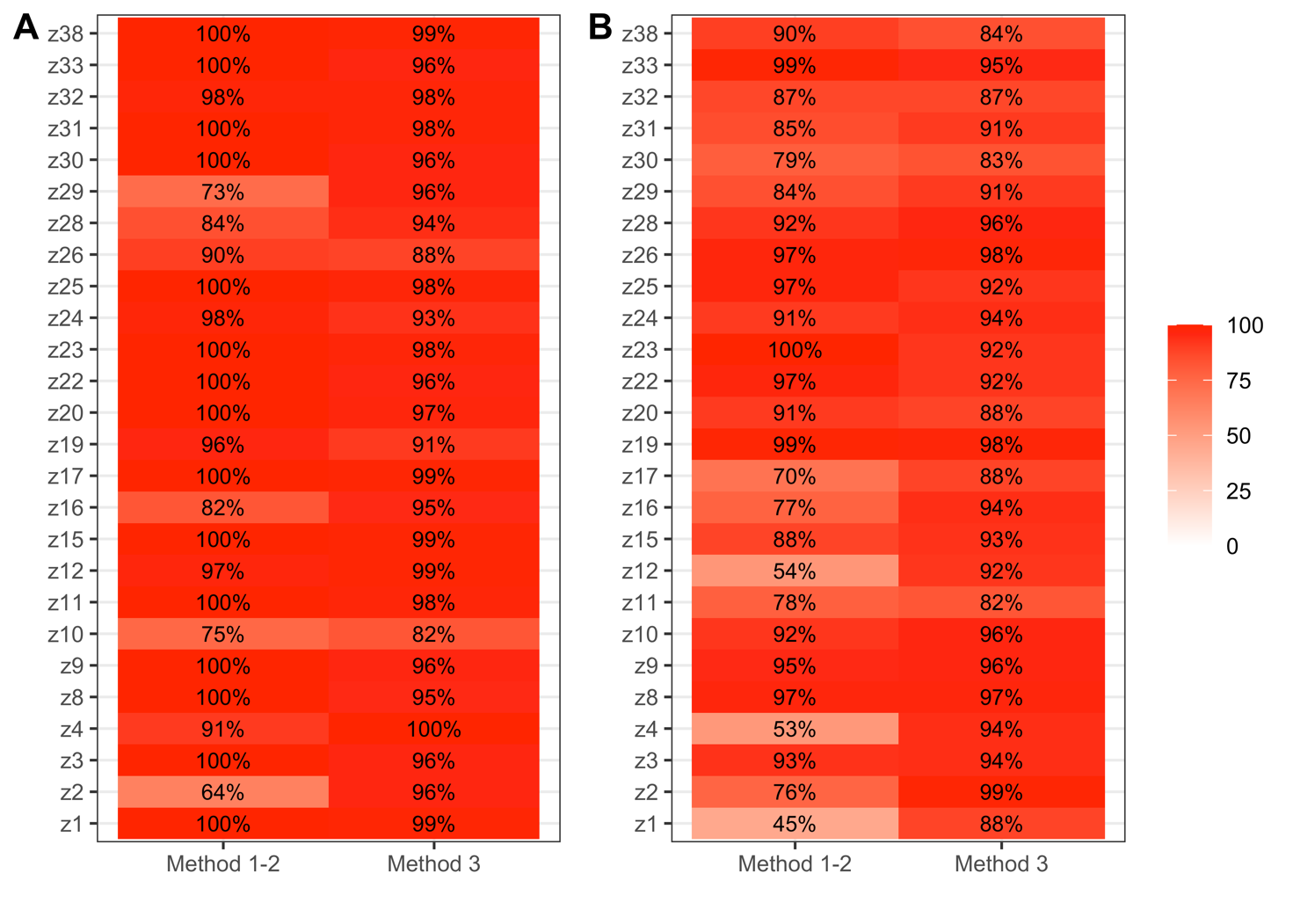

**Figure S6**: Box-plots of the bias in the estimates of the regression parameter associated to the single WQS index of the two methods.

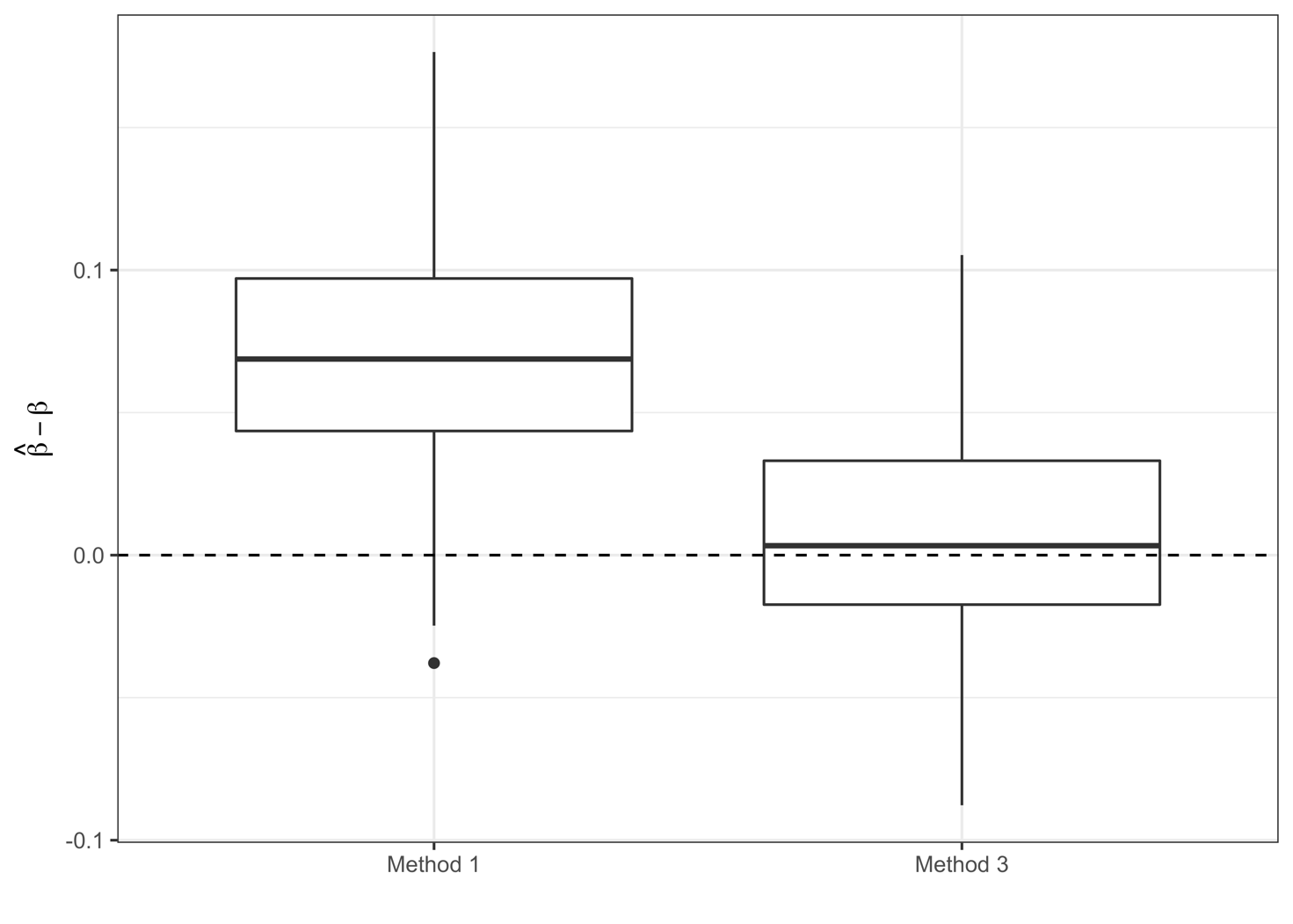

**Figure S7**: Heatmap of the sensitivity (panel A) and specificity (panel B) of the two methods in detecting the elements with a weight greater than 0 and those with a null weight when considering a single index.

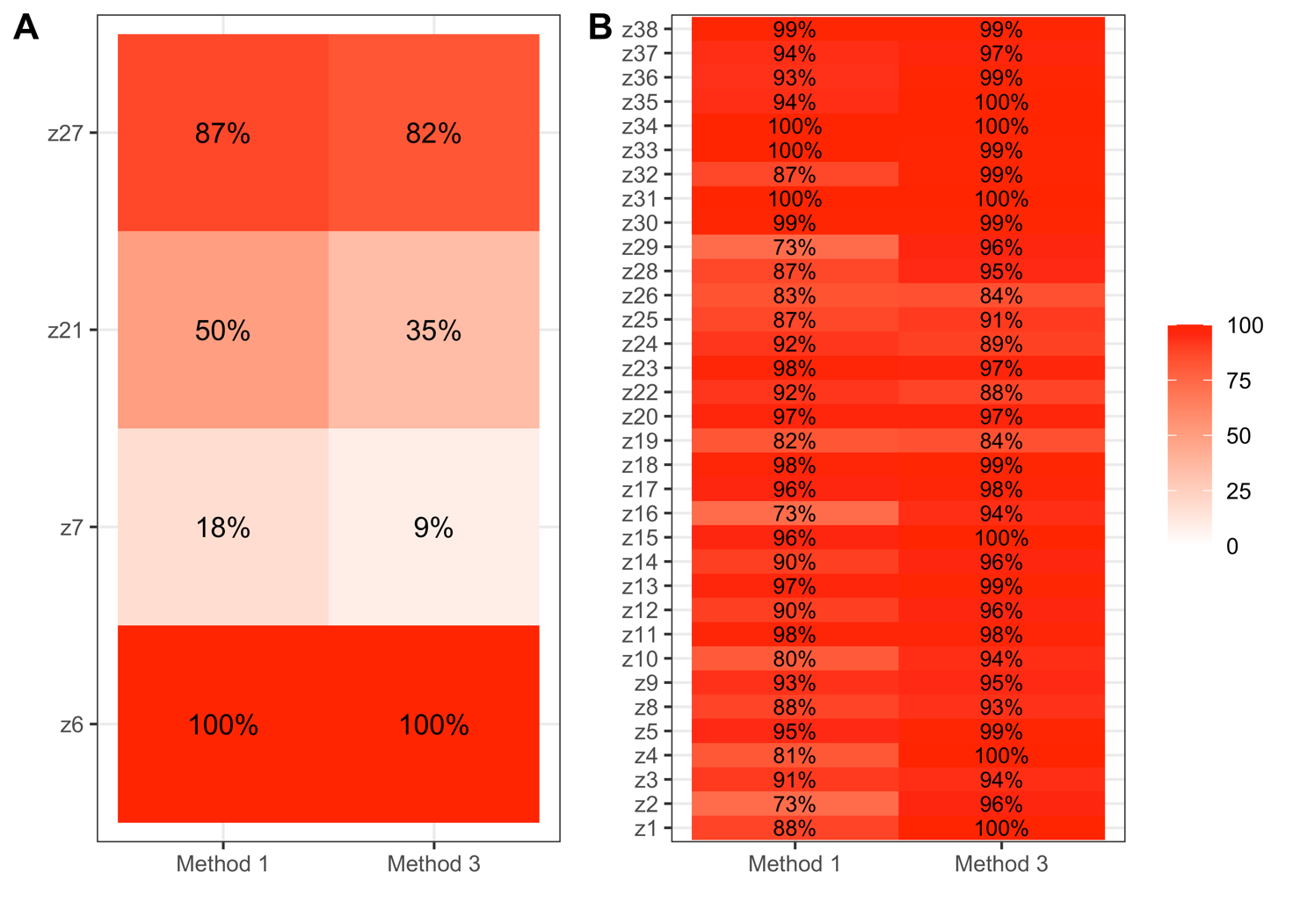
